# Associations between prenatal exposure to gestational diabetes mellitus and child adiposity markers: mediating effects of brain structure

**DOI:** 10.1101/2022.02.03.22270398

**Authors:** Shan Luo, Eustace Hsu, Katherine E. Lawrence, Shana Adise, Trevor A. Pickering, Megan M. Herting, Thomas Buchanan, Kathleen A. Page, Paul M. Thompson

## Abstract

**Objectives:** To investigate neural mechanisms underlying increased obesity risk in children prenatally exposed to gestational diabetes mellitus (GDM).

**Research Design and Methods:** This was a cross-sectional study of 9-10-year-old participants and siblings across the United States. Data was obtained from the baseline assessment of the Adolescent Brain and Cognitive Development (ABCD) Study® in which brain structure was evaluated by magnetic resonance imaging. Adiposity markers included age- and sex-specific body mass index (BMI *z-*scores), waist circumference and waist-to-height ratio. GDM exposure was self-reported, and discordance for GDM exposure within biological siblings was identified. Mixed effects and mediation models were used to examine associations between prenatal GDM exposure, brain structure, and adiposity markers with sociodemographic covariates.

**Results:** The sample included 8,521 children (age: 9.9±0.6 years; 51% males; 7% GDM-exposed), among whom there were 28 sibling pairs discordant for GDM exposure. Across the entire study sample, prenatal exposure to GDM was associated with lower global and regional cortical gray matter volume (GMV) in the bilateral rostral middle frontal gyrus and superior temporal gyrus. In a subset of sample only including siblings, GDM-exposed siblings also demonstrated lower global cortical GMV than un-exposed siblings. Global cortical GMV partially mediated the associations between prenatal GDM exposure and child adiposity markers.

**Conclusions:** Our results establish robust and generalizable brain markers of prenatal GDM exposure and suggest that low cortical GMV may explain increased obesity risk for offspring prenatally exposed to GDM.

**Twitter summary:** Prenatal exposure to gestational diabetes mellitus #GDM is associated with smaller cortical gray matter volume #brain, which in turn relates to larger adiposity markers #BMI in a large and diverse cohort of 8521 children.

**Article Highlights:**

- Prenatal exposure to gestational diabetes mellitus (GDM) is associated with smaller cortical gray matter volume (GMV) in a large and diverse cohort of 8521 children, independent from genetics and shared environment.
- Cortical GMV partially mediated the associations of prenatal GDM exposure and adiposity markers in children.
- These results establish robust and generalizable brain markers of prenatal GDM exposure and provide neurobiological underpinnings of increased obesity risk in offspring prenatally exposed to GDM.

## Introduction

One in five youth in the U.S. are living with obesity(1), placing them at increased risk for diabetes and cardiovascular disease(2). Reducing the incidence of childhood obesity would have a consequential impact on the individual and socioeconomic burdens associated with obesity, however existing strategies for treating adult obesity do not achieve long-lasting weight loss in this younger population(3–6) and more than 90% of youth with obesity will continue to be classified as overweight or obese in adulthood(7, 8). Improved understanding of risk factors for pediatric obesity will advance our knowledge of how obesity develops and guide interventions to mitigate short- and long-term disease risks.

Offspring exposed to GDM prenatally have an increased risk of developing pediatric obesity(9–15), a larger body mass index (BMI) measures, waist-to-hip circumference, and waist-to-height ratio (WHtR) (14, 16–18). Furthermore, the BMI of offspring exposed to GDM was greater than that of their siblings who experienced nondiabetic pregnancies(19, 20), suggesting that the effect of prenatal GDM exposure on offspring BMI is independent of shared environment and genetics. The high prevalence and persistence of childhood obesity highlights the importance of understanding the underlying mechanisms.

While the mechanisms for increased obesity risk in GDM-exposed offspring are unknown, existing studies suggest a neural basis. Serum levels of neurotrophins including nerve growth factor (NGF) and brain-derived neurotrophic factor (BDNF) – essential for neuronal growth, development, and differentiation – are lower in pregnant women with diabetes(21) and infants prenatally exposed to GDM(22). Offspring exposed to GDM prenatally demonstrated brain structural alterations, such as presence of hypothalamic gliosis, lower white matter integrity in sensorimotor regions(23), lower cortical excitability(24), and mixed results on the hippocampal thickness(25, 26). However, this work is hampered by small sample sizes and poor representation of diverse populations, calling into question the reproducibility and generalizability of findings. Prior studies are also limited in their ability to establish mediating relationships between prenatal exposure to GDM, brain and adiposity in children. Moreover, it remains to be determined whether neural correlates of prenatal GDM exposure are independent of genetics and shared environment.

To elucidate gaps in the literature, we leveraged brain and anthropometric data in children aged 9 to 10 years old from the Adolescent Brain and Cognitive Development (ABCD) Study® - the largest and most diverse pediatric neuroimaging study to date(27). We examined relationships between prenatal exposure to GDM and brain structural measures. Furthermore, a mediation analysis was performed to investigate whether brain structural measurements mediated associations between prenatal GDM exposure and adiposity markers in children. The ABCD study also includes siblings discordant for GDM exposure providing the opportunity to control for genetics and shared environment. We hypothesized that GDM-exposed children (vs. un-exposed children) would have lower brain structural measures, independent of genetics and shared environment; brain structural measures would partially mediate associations of GDM exposure with adiposity markers in children.

## Research Design and Methods

### Participants

ABCD is a large-scale, 10-yr longitudinal study of pediatric rain and cognitive development in the US. Participants and siblings were initially recruited from a randomized sample of schools within range of 21 sites across four major regions of the U.S. (Northeast, Midwest, South, West)(28). Recruitment was optimized to accurately represent the sociodemographic diversity of the U.S. Institutional Review Board (IRB) approval was obtained from the centralized IRB at the University of California, San Diego (IRB# 160091), as well as at each data collection site.

Data for this project was obtained from the ABCD 3.0 data release (DOI: 10.15154/1524739) and focused on baseline assessments collected from 9/1/2016 to 10/15/2018. Participants were excluded if they met the following criteria: not fluent in English, history of seizures, birth more than 12 weeks premature, birth weight less than 1200 grams, complications at birth, substance use disorder, intellectual disability, traumatic brain injury, brain tumor, stroke, aneurysm, brain hemorrhage, subdural hematoma, cerebral palsy, diabetes, lead poisoning, muscular dystrophy, autism spectrum disorder (ASD), and other medical conditions considered exclusionary(29). Additional exclusion criteria were applied based on anthropometric measurements, neuroimaging, and other covariates as described in the following sections, resulting in a final sample of 8,521 participants (eFigure 1). Briefly, subjects were only included if their anthropometric and neuroimaging measurements passed quality control and if they had complete data for additional covariates.

### Gestational diabetes mellitus exposure

GDM exposure was self-reported by the parent during baseline assessment via a question: “During the pregnancy with this child, did you/biological mother have pregnancy-related diabetes?”.

Biological siblings with discordance for GDM exposure were restricted to participants who shared the same mother. Birth order was calculated based on interview age and interview date.

### Anthropometric measures

Children’s weight, height, and waist circumference were measured at baseline by a trained researcher(30). Waist circumference was measured with a tape around the highest point on the pelvic bone. Height (in inches) and weight (in pounds) were recorded as the mean of up to three separate measurements. These measures were used to calculate WHtR and BMI (kg/m^2^).

Age- and sex-specific BMI percentiles, BMI *z*-scores, weight *z*-scores and height *z*-scores were calculated according to the Center for Disease Control (CDC) guidelines(31, 32). Childhood weight status is defined using CDC guidelines(33): obesity (≥95^th^ %ile), overweight (≥ 85^th^ %ile to <95^th^ %ile), normal weight (≥5^th^ %ile to < 85^th^ %ile) and underweight (<5^th^ %ile). Waist *z*-scores were calculated based on NHANES III data. Baseline anthropometrics data were excluded based on the following(34, 35): 1) BMI *z*-scores ≤ -4 SDs or ≥8 SDs, 2) BMI <10, 3) weight *z*-scores ≤-5 SDs or ≥8 SDs, 4) height *z*-scores or waist *z*-scores <-4 SDs or >4 SDs, 5) WHtR ≤0.2 or ≥1.

### Neuroimaging

Magnetic resonance imaging (MRI) data collection methods were optimized for 28 3-T scanners across 21 ABCD Study sites(36). FreeSurfer (version 5.3.0) was used for cortical surface reconstruction and subcortical segmentation, based on the T1-weighted anatomical scans, including cortical gray and white matter as well as subcortical volumes (mm^3^) using automated segmentation, and regional cortical thickness (mm), and regional cortical surface area (mm^2^) using the Desikan-Killiany Atlas(37, 38).

Neuroimaging analyses excluded participants who had abnormal radiological findings or whose T1 scan was of insufficient quality, as determined by the ABCD Data Analytics and Informatics Center(36). Participants were also excluded based on quality control procedures on the cortical surface reconstruction for five categories of inaccuracy: severity of motion, intensity inhomogeneity, white matter underestimation, pial overestimation, and magnetic susceptibility artifact(36).

### Covariates

Race/ethnicity was categorized into five groups: Hispanic, non-Hispanic White, non-Hispanic Black, non-Hispanic Asian, and Other (see Garavan et al., 2018(29), for further details). Parental education was modeled as a binary variable indicating whether at least one parent has obtained a bachelor’s degree. Yearly household income was quantified as an ordinal range which included three categories: < $50,000, ≥$50,000 and <$100,000, and ≥$100,000. Pubertal stage was calculated using the parent-report pubertal development scale and reduced to three levels, pre-pubertal, early pubertal, and mid-to post-pubertal stage.

### Data Analysis

Linear mixed effects models were used to examine associations of GDM exposure with global brain measurements and adiposity markers. Global brain measurements included total cortical surface area, mean cortical thickness, total cortical gray matter volume (GMV), subcortical GMV, and cerebral white matter volume. Adiposity measures included BMI *z*-scores, waist circumference and WHtR.

Two sets of models were fit. The first set of models included the full sample. Family ID, nested within the site, were modeled as random intercepts to account for shared variation related to study visit location and family membership. Covariates included sociodemographic variables such as age, sex assigned at birth, pubertal status, race/ethnicity, family income, and parental education. Age and sex were not included as covariates for models with BMI *z*-score as a dependent variable. Imaging analyses also included scanner model, handedness, and intracranial volume (ICV) for volumetric measures. Additional models were fitted to adjust for: 1) gestational age at birth, 2) maternal other health problems during pregnancy, 3) child health problems at birth, 4) maternal alcohol or tobacco use during pregnancy. To follow up with significant associations of GDM exposure with global brain measures, we conducted region-of-interest (ROI) analysis.

Mediation analysis was completed with the hypothesis that the correlation between prenatal GDM exposure and adiposity markers was mediated by brain structural measurements, using the *mediation* package in R(39). First, confounding covariates (i.e., sociodemographic variables, scanner, handedness, and ICV) were regressed out of adiposity and brain structure variables using regressions. Next, separate components of the possible mediation paths were assessed (A: correlation between GDM exposure and brain; B: correlation between GDM exposure and adiposity; C: correlation between brain and adiposity in a model that also includes GDM exposure)(40). Mediation was then tested in models where all component paths were significant. Nonparametric bootstrapped percentile confidence intervals were estimated from 1,000 simulations.

To further study if brain correlates of GDM exposure identified from the full sample are independent of genetics and shared environment, we performed a second set of model fits only in sibling pairs discordant for GDM exposure. Models included family ID as a random intercept and adjusted for birth order, sex, race/ethnicity, pubertal status, family income, parental education, handedness, and ICV for volumetric analyses. Site and scanner were excluded due to insufficient sample size. Age was not adjusted for as it was highly correlated with birth order in this sample.

Analyses used linear mixed effects models in R with the *lme4* package. Standardized betas were reported. 95% Wald confidence intervals (CI) were calculated based on the local curvature of the likelihood surface. Cohen’s *d* effect sizes were calculated from least-squares means, model residual standard deviation, and residual degrees-of-freedom using the *emmeans* package. *P*-values were calculated using Satterthwaite’s method in the *lmerTest* package. Tests of significance (2-tailed) were corrected for multiple comparisons using the Benjamini-Hochberg false discovery rate (FDR) correction, with *P*<0.05 as the corrected threshold for significance.

### Data and Resource Availability

The datasets generated during and/or analyzed in the current study are available from the corresponding author upon reasonable request.

## Results

### Participant Demographics

Characteristics of the sample are described in **Table 1**. Around 7% of children were prenatally exposed to GDM and differed from un-exposed children in terms of race/ethnicity (Χ^2^(4) = 29.505, *P*<0.001), family income (Χ^2^(2) = 11.728, *P*=0.003), parental education (Χ^2^(1) = 16.988, *P*<0.001), and pubertal status (Χ^2^(2) = 10.960, *P*=0.028).

**Table 1.**
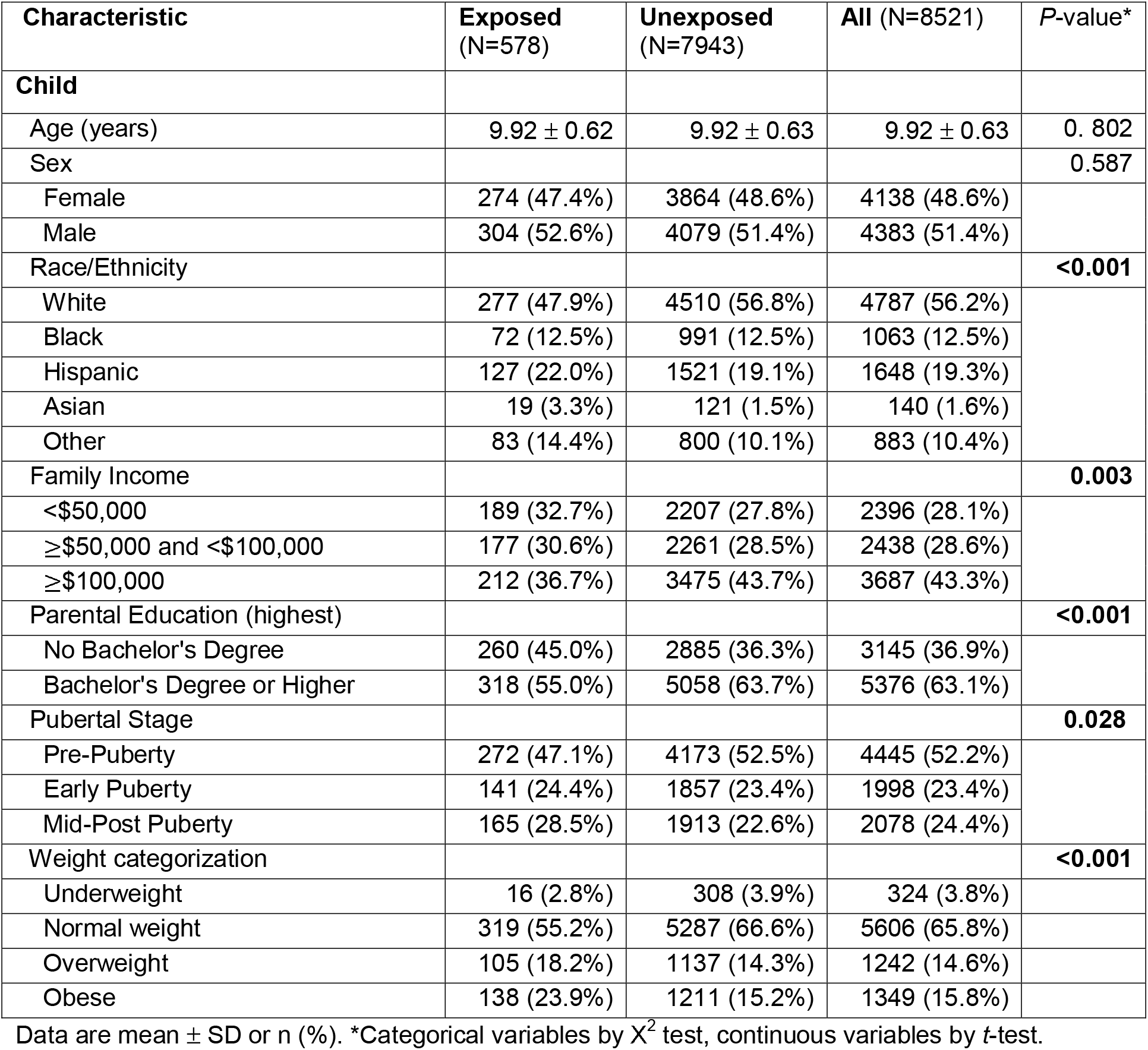
Sample Characteristics by Gestational Diabetes Mellitus Status.

| Characteristic | Exposed<br>(N=578) | Unexposed<br>(N=7943) | All (N=8521) | P-value* |
| --- | --- | --- | --- | --- |
| **Child** |  |  |  |  |
| Age (years) | 9.92 ± 0.62 | 9.92 ± 0.63 | 9.92 ± 0.63 | 0.802 |
| Sex |  |  |  | 0.587 |
| Female | 274 (47.4%) | 3864 (48.6%) | 4138 (48.6%) |  |
| Male | 304 (52.6%) | 4079 (51.4%) | 4383 (51.4%) |  |
| Race/Ethnicity |  |  |  | **<0.001** |
| White | 277 (47.9%) | 4510 (56.8%) | 4787 (56.2%) |  |
| Black | 72 (12.5%) | 991 (12.5%) | 1063 (12.5%) |  |
| Hispanic | 127 (22.0%) | 1521 (19.1%) | 1648 (19.3%) |  |
| Asian | 19 (3.3%) | 121 (1.5%) | 140 (1.6%) |  |
| Other | 83 (14.4%) | 800 (10.1%) | 883 (10.4%) |  |
| Family Income |  |  |  | **0.003** |
| <\$50,000 | 189 (32.7%) | 2207 (27.8%) | 2396 (28.1%) | |
| ≥\$50,000 and <\$100,000 | 177 (30.6%) | 2261 (28.5%) | 2438 (28.6%) | |
| ≥\$100,000 | 212 (36.7%) | 3475 (43.7%) | 3687 (43.3%) | |
| Parental Education (highest) |  |  |  | **<0.001** |
| No Bachelor's Degree | 260 (45.0%) | 2885 (36.3%) | 3145 (36.9%) |  |
| Bachelor's Degree or Higher | 318 (55.0%) | 5058 (63.7%) | 5376 (63.1%) |  |
| Pubertal Stage |  |  |  | **0.028** |
| Pre-Puberty | 272 (47.1%) | 4173 (52.5%) | 4445 (52.2%) |  |
| Early Puberty | 141 (24.4%) | 1857 (23.4%) | 1998 (23.4%) |  |
| Mid-Post Puberty | 165 (28.5%) | 1913 (22.6%) | 2078 (24.4%) |  |
| Weight categorization |  |  |  | **<0.001** |
| Underweight | 16 (2.8%) | 308 (3.9%) | 324 (3.8%) |  |
| Normal weight | 319 (55.2%) | 5287 (66.6%) | 5606 (65.8%) |  |
| Overweight | 105 (18.2%) | 1137 (14.3%) | 1242 (14.6%) |  |
| Obese | 138 (23.9%) | 1211 (15.2%) | 1349 (15.8%) |  |

Data are mean ± SD or n (%). \*Categorical variables by $\chi^2$ test, continuous variables by *t*-test.

Among sibling pairs discordant for GDM exposure, higher rates of GDM exposure were found in the older siblings than younger ones (71% vs 29%, Χ^2^(1) = 8.643, *P*=0.003). Discordant siblings for GDM exposure did not differ in any other covariates.

### GDM Exposure and Child Global and Regional Brain Measurements

Prenatal exposure to GDM was associated with smaller brain structure across global brain measures in the full sample (**Figure 1A, Table 2**). Effect sizes (Cohen’s *d*) for GDM-exposed vs. un-exposed children in global brain measures ranged from -0.089 to -0.160. Only total cortical GMV remained significant after FDR correction (β (95% CI) = -0.051 (−0.089, -0.013), FDR corrected *P*=0.045). Total cortical surface area and subcortical GMV were trended towards significance after FDR correction. Unadjusted results are included in eTable 1. In a subset of sample with sibling pairs discordant for GDM exposure, total cortical GMV was also lower in GDM-exposed siblings than un-exposed siblings (β (95% CI) = -0.284 (−0.531, -0.037), *P*=0.034, **Figure 1A**).

**Figure 1.** Relationships between exposure to gestational diabetes mellitus (GDM) and child brain structure. **A**). Bar plots display mean ± 95% CI of adjusted cortical gray matter volume (GMV), separated by prenatal GDM exposure status. Statistics for full sample adjusted for family ID nested within site, scanner model, handedness, age, sex, pubertal status, race/ethnicity, family income, parental education, and statistics for subsample of siblings discordant in GDM exposure adjusted for family ID, handedness, birth order, sex, pubertal status, race/ethnicity, family income, parental education. **B**). Associations between GDM exposure and child regional cortical gray matter volume in the significant regions of interest. T score denotes regions significantly associated with GDM exposure from linear mixed effects models (FDR corrected *P*<0.05) where family ID nested within site, as well as subject, were included as random effects, and age, sex, pubertal status, race/ethnicity, family income, parental education, scanner model, intracranial volume, and handedness were included as fixed effects. All regions of interest analysis were bilateral in the models.

**Table 2.** Associations between Gestational Diabetes Mellitus Exposure and Global and Regional Brain Measurements.

| Model | $\beta^*$ | 95% CI** | P-value | FDR-adjusted P-value*** | Cohen's *d* |
| --- | --- | --- | --- | --- | --- |
| **Global Brain Measurements** |  |  |  |  |  |
| Total Cortical Surface Area(mm<sup>2</sup>) | -0.073 | (-0.144, -0.003) | 0.042 | 0.069 | -0.144 |
| Mean Cortical Thickness (mm) | -0.057 | (-0.132, 0.017) | 0.132 | 0.132 | -0.089 |
| Cortical Gray Matter Volume(mm<sup>3</sup>) | -0.051 | (-0.089, -0.013) | 0.009 | **0.045** | -0.160 |
| Subcortical Gray Matter Volume(mm<sup>3</sup>) | -0.048 | (-0.095, -0.001) | 0.044 | 0.074 | -0.128 |
| Cerebral White Matter Volume(mm<sup>3</sup>) | -0.024 | (-0.062, 0.014) | 0.220 | 0.220 | -0.079 |
| **Regional Cortical Gray Matter Volume (mm<sup>3</sup>)** |  |  |  |  |  |
| Banks of the Superior Temporal Sulcus | -0.037 | (-0.097, 0.022) | 0.217 | 0.393 | -0.048 |
| Caudal Anterior Cingulate Cortex | 0.003 | (-0.055, 0.060) | 0.924 | 0.924 | 0.003 |
| Caudal Middle Frontal Gyrus | -0.016 | (-0.077, 0.045) | 0.607 | 0.765 | -0.026 |
| Cuneus Cortex | -0.083 | (-0.145, -0.021) | 0.009 | 0.097 | -0.130 |
| Entorhinal Cortex | 0.059 | (-0.004, 0.122) | 0.066 | 0.250 | 0.078 |
| Fusiform Gyrus | -0.029 | (-0.086, 0.027) | 0.305 | 0.494 | -0.050 |
| Inferior Parietal Cortex | -0.032 | (-0.089, 0.026) | 0.284 | 0.482 | -0.057 |
| Inferior Temporal Gyrus | 0.036 | (-0.020, 0.093) | 0.209 | 0.393 | 0.064 |
| Isthmus Cingulate Cortex | -0.012 | (-0.071, 0.047) | 0.681 | 0.772 | -0.019 |
| Lateral Occipital Cortex | -0.052 | (-0.107, -0.004) | 0.067 | 0.250 | -0.098 |
| Lateral Orbital Frontal Cortex | -0.044 | (-0.099, -0.011) | 0.116 | 0.328 | -0.092 |
| Lingual Gyrus | -0.058 | (-0.124, 0.007) | 0.082 | 0.253 | -0.115 |
| Medial Orbital Frontal Cortex | -0.012 | (-0.068, 0.044) | 0.678 | 0.772 | -0.018 |
| Middle Temporal Gyrus | -0.040 | (-0.094, -0.014) | 0.146 | 0.354 | -0.079 |
| Parahippocampal Gyrus | 0.025 | (-0.038, 0.087) | 0.440 | 0.676 | 0.034 |
| Paracentral Lobule | -0.040 | (-0.102, 0.022) | 0.211 | 0.393 | -0.055 |
| Pars Opercularis | -0.018 | (-0.080, 0.044) | 0.566 | 0.740 | -0.025 |
| Pars Orbitalis | -0.055 | (-0.116, 0.005) | 0.074 | 0.250 | -0.079 |
| Pars Triangularis | -0.020 | (-0.086, 0.047) | 0.047 | 0.560 | -0.028 |
| Pericalcarine Cortex | -0.089 | (-0.159, -0.020) | 0.118 | 0.100 | -0.181 |
| Postcentral Gyrus | -0.037 | (-0.096, 0.022) | 0.220 | 0.393 | -0.066 |
| Posterior Cingulate Cortex | 0.008 | (-0.066, 0.110) | 0.620 | 0.787 | 0.011 |
| Precentral Gyrus | 0.020 | (-0.035, 0.075) | 0.477 | 0.677 | 0.038 |
| Precuneus Cortex | -0.035 | (-0.090, 0.020) | 0.216 | 0.393 | -0.078 |
| Rostral Anterior Cingulate Cortex | -0.013 | (-0.070, 0.044) | 0.654 | 0.772 | -0.017 |
| Rostral Middle Frontal Gyrus | -0.087 | (-0.143, -0.031) | 0.002 | **0.042** | -0.179 |
| Superior Frontal Gyrus | -0.054 | (-0.106, -0.002) | 0.042 | 0.0236 | -0.120 |
| Superior Parietal Cortex | -0.044 | (-0.104, 0.015) | 0.146 | 0.354 | -0.086 |
| Superior Temporal Gyrus | -0.098 | (-0.154, -0.043) | <0.001 | **0.017** | -0.195 |
| Supramarginal Gyrus | -0.009 | (-0.065, 0.047) | 0.748 | 0.820 | -0.013 |
| Frontal Pole | -0.006 | (-0.069, 0.057) | 0.848 | 0.873 | -0.008 |
| Temporal Pole | 0.024 | (-0.042, 0.090) | 0.472 | 0.676 | -0.031 |
| Transverse Temporal Cortex | -0.077 | (-0.141, -0.013) | 0.019 | 0.130 | -0.113 |
| Insular Cortex | -0.051 | (-0.106, 0.004) | 0.069 | 0.250 | -0.125 |

\* A positive $\beta$ corresponds to larger measurements in the GDM-exposed group, whereas a negative $\beta$ corresponds to the opposite. \*\* Regression coefficient (95% CI) from linear mixed effects models. For regional cortical measurements, coefficient (95% CI) represents main effect estimate of prenatal exposure to GDM. GDM-exposure by hemisphere interactions were modeled but not significant in any region of interest. \*\*\* Multiple comparisons are conducted with Benjamini-Hochberg FDR correction. Boldface indicates significance at the corrected threshold of $P < 0.05$ .

ROI analysis (**Table 2, Figure 1B)** found that prenatal exposure to GDM was associated with smaller cortical GMV in the bilateral rostral middle frontal gyrus (MFG) (β (95% CI) = -0.087 (−0.143, -0.031), *P*=0.042) and superior temporal gyrus (STG) (β (95% CI) = -0.098 (−0.154, -0.043), *P*= 0.017). The effect size for GDM-exposed vs. un-exposed children in the MFG and STG was -0.179 and -0.195 respectively. Results for global and regional cortical GMV remained largely the same after additional adjustment for gestational age, other maternal health problems during pregnancy, child health problems at birth, or maternal alcohol/tobacco use during pregnancy (eTable 2-9).

Given prior studies reported conflicting results for associations of prenatal GDM exposure and hippocampal GMV, for completeness we present ROI analysis results for subcortical regions in the Supplement (eTable 10). There were no significant associations between prenatal GDM exposure and GMV in any subcortical ROIs.

### GDM Exposure and Child Adiposity Markers

GDM exposure was associated with a higher likelihood of being classified as overweight or obese (42 vs 29%: Χ^2^(3) = 43.160, *P*<0.001). Children exposed to GDM (vs. un-exposed children) had greater adiposity markers (BMI *z*-scores: β (95% CI) = 0.175 (0.093, 0.256), FDR corrected *P*<0.001; Waist circumference: β (95% CI) = 0.201 (0.121, 0.281), FDR corrected *P*<0.001; Waist-to-height ratio (WHtR): β (95% CI) = 0.199 (0.119, 0.280), FDR corrected *P*<0.001) (eFigure 2).

### Mediation Relationships among GDM Exposure, Brain, and Adiposity Markers

Global cortical GMV partially mediated the relationship between GDM exposure and BMI *z*-scores, with a significant indirect effect (mediated effect (95% CI) = 0.007 (0.002, 0.096), *P*=0.012). The direct effect of GDM on BMI *z*-score was reduced from β (95% CI) = 0.168 (0.077, 0.255), *P*<0.001 to β (95% CI) = 0.161 (0.072, 0.247), *P*<0.001 in the presence of the mediator (**Figure 2**), indicating a partial mediation. Global cortical GMV also partially mediated the relationships between GDM exposure and waist circumference (mediated effect (95% CI) = 0.008 (0.002, 0.015), *P*=0.012), and WHtR (mediated effect (95% CI) = 0.005 (0.001, 0.009), *P*=0.012) (eTable 1).

**Figure 2.** Cortical gray matter volume partially mediating the relationships between GDM exposure and BMI *z*-scores. Variation based on pubertal status, race/ethnicity, family income, and parental education was regressed out of BMI z-scores, and additional variation based on scanner, intracranial volume, and handedness was regressed out of cortical gray matter volume prior to conducting mediation modeling. Reported values for Paths A, B, C and C’ indicate standardized regression coefficients for each path. Paths B and C’ indicate standardized multiple regression coefficients of GDM exposure (C’) and cortical GMV (B) in predicting BMI *z*-scores. The mediated effect indicates the reduced direct effect of GDM exposure on BMI *z*-scores when the mediator is included in this model. * *P* < 0.05, *** *P* < 0.001

### Conclusions

This is the first large, multi-site study investigating relationships between prenatal exposure to GDM, brain structure, and adiposity markers in a diverse cohort of 8521 children. We found that prenatal exposure to GDM was associated with lower global cortical GMV in the entire study sample as well as a subset of sample including siblings discordant for GDM exposure. And global cortical GMV in part mediated relationships between prenatal GDM exposure and adiposity markers in children. The results established robust and generalizable brain signatures of prenatal GDM exposure and provided a neurobiological pathway by which prenatal exposure to GDM increases obesity risk in offspring.

A link between prenatal GDM exposure and altered neural structure has been previously described (18, 23, 25, 41–43); however interpretation has been constrained by methodological limitations including small sample sizes, poor representation of diverse populations and lack of controlling for genetics and shared environment. Capitalizing upon the large and diverse sample of the ABCD study®, we were able to establish robust and generalizable neural correlates of prenatal GDM exposure, including lower global cortical GMV in youth aged between 9 and 10 years. Incorporating covariates known influence brain development(44–47), including gestational age, other maternal health problems during pregnancy (e.g., high blood pressure, pre-eclampsia), child health problems at birth, and maternal alcohol /tobacco use during pregnancy did not impact the results. Although the effect observed are small, they are nonetheless comparable with other changes observed in the context of brain disorders including attention deficit hyperactivity disorder, ASD, obsessive compulsive disorder, and major depressive disorder(48–50). Data from sibling pairs discordant for GDM exposure also demonstrated lower total cortical GMV in GDM-exposed siblings than un-exposed siblings, implying the effect of prenatal GDM exposure on cortical GMV is likely via intrauterine mechanism since sibling design controlled for genetics and shared environment.

While alterations in brain development are hypothesized as a potential mechanism for obesity risk in GDM-exposed offspring, this hypothesis has not been fully tested in humans. Mediation models found that global cortical GMV partially mediated the association between prenatal exposure to GDM and adiposity markers in children. These findings expand on prior studies(18, 42) by showing direct evidence of the mediating role of the brain in the associations of prenatal exposure to GDM and offspring adiposity, and provide a neurobiological explanation of increased obesity risk in GDM-exposed offspring. Given the cross-sectional nature of ABCD baseline data, causality cannot be established. As well, we cannot exclude other possible mediating relationships between prenatal GDM exposure, brain, and adiposity in children.

Our analysis identified a selective impact of prenatal GDM exposure upon the MFG and STG. The frontal and temporal regions follow a protracted developmental trajectory, an inverted U-shape trajectory with a pre-adolescent increase followed by post-adolescent decrease(51), which render them particularly vulnerable to early insult, such as that inflicted by GDM exposure. A recent study using the ABCD dataset reported that lower GMV in the MFG is associated with greater concurrent BMI *z*-scores and greater changes in BMI *z*-scores in one year(52). Together, these results suggest that GDM-induced changes in cortical GMV may be associated with both current and future weight status. While the functional significance of GDM exposure-associated alterations in cortical GMV remains to be established, we can reasonably speculate that functional deficits in cortical areas implicated in dietary self-regulation may lead to greater adiposity in children exposed to GDM given that 1) cortical regions, particularly the MFG, play a critical role in self-regulation, including top down regulation of craving and food consumption(53, 54); and 2) smaller cortical GMV in prefrontal areas has been shown to be associated with poorer self-regulation(55), which is tightly linked to overeating and obesity(56). Additional studies using the ABCD dataset found other neural correlates of obesity or weight gain, including lower cortical thickness (primarily in the prefrontal cortex)(57) and higher cell density in the nucleus accumbens(58). We recognize that GDM exposure is one of many factors contributing to childhood obesity, thus other obesity-associated risk factors may be contributing to the changes in neural structure. In contrast to animal studies(59–61) and a recent human study(25), we and others (26)did not identify a significant association of GDM exposure with hippocampal GMV in children. We speculate that this inconsistency may be due to assessment of specific sub-regions of the hippocampus by former studies vs. whole hippocampus by latter studies.

Our findings have important clinical implications. This was a well-powered study with over 8,500 children from different demographic and socio-economic backgrounds, demonstrating that there were adverse effects of prenatal exposure to GDM on brain development in children. It is important for clinicians to be aware of detrimental effects of diabetes during pregnancy on the developing brain in offspring. Future study should examine potential interventions that may mitigate adverse effects of prenatal GDM exposure on offspring brain development, thereby reducing obesity risk.

Our study also includes some limitations. GDM exposure was self-reported; however, the reported prevalence in the ABCD study was comparable with estimated rate of GDM in the US. Detailed information on treatment and severity of diabetes in pregnancy were not available; thus, we were not able to assess relationships between those variables and child brain structure. The ABCD study® did not record maternal obesity status during pregnancy, which is closely related to maternal diabetes. Our prior work showed overlapping and differential effects of maternal obesity exposure and GDM exposure on children’s brain metabolic and reward circuitry(18, 42, 62). Thus, we speculate that maternal obesity exposure may have similar or separate effects from those of maternal diabetes in global and regional cortical GMV.

In conclusion, prenatal GDM exposure is associated with smaller cortical GMV, independent from genetics and shared environment, which is in turn related to greater markers of adiposity in children. Low cortical GMV may be a potential neural mechanism by which prenatal GDM exposure mediates obesity risk in offspring.

## Data Availability

All data produced are available online at NIMH data archive.

## Funding Assistance

This work was supported by the National Institutes of Health (NIH) National Institute of Diabetes and Digestive and Kidney Diseases K01DK115638 (PI: SL), R03DK129186 (PI: SL) and USC Diabetes and Obesity Research Institute Pilot Award (PI: SL). This study was partially supported by NIMH F32MH12205 (PI: KL).

## Author Contributions

S.L. and E.H. performed the statistical analysis and drafted the manuscript. All authors provided review, commentary, and revisions to the manuscript, approved the final manuscript as submitted. S.L. is the guarantor of this work and, as such, had full access to all the data in the study and takes responsibility for the integrity of the data and the accuracy of the data analysis.

## Prior Presentation

Parts of this study were presented as a poster at the annual meeting of the Organization for Human Brain Mapping, Glasgow, Scotland, 19-23 June 2022.

