## Supplemental Materials for "Associations between prenatal exposure to gestational diabetes mellitus and child adiposity markers: mediating effects of brain structure"

**Supporting Information Text**

**eMethods**

*Participant Exclusion*

Details regarding inclusion of participants are described in the main text and eFigure 1. Additional analyses excluded participants based on missing values for variables added to analysis: gestational age, health problems during pregnancy, health problems at birth, alcohol/tobacco use during pregnancy, and area deprivation index~~.~~

*Additional Covariates*

*Gestational Age*

Parents self-reported whether the child was born premature and, if so, how many weeks premature. Gestational age was derived as 40 weeks for children not born premature, and otherwise by subtracting the number of weeks premature from 40.

*Health Problems During Pregnancy*

Maternal health problems during pregnancy was modeled as a binary variable indicating a self-reported response to any of the options to the question “During the pregnancy with the child, did you/biological mother have any of the following conditions?,” with possible options being considered as: 1) severe nausea or vomiting extending past the sixth month or accompanied by weight loss, 2) heavy bleeding requiring bed rest or special treatment, 3) pre-eclampsia, eclampsia or toxemia, 4) severe gall bladder attack, 5) persistent proteinurea, 6) rubella during first three months of pregnancy, 7) severe anemia, 8) urinary tract infections, 9) pregnancy-related high blood pressure, 10) previa, abruptio, or other problems with the placenta, 11) an accident or injury requiring medical care. The response to an additional option, pregnancy-related diabetes, was separately assessed as the main predictor of interest.

*Health Problems at Birth*

Infant’s health problem(s) at birth was modeled as a binary variable indicating a parental response to any option for the question “Did he/she have any of the following complications at birth?” with possible options as: 1) blue at birth, 2) slow heartbeat, 3) did not breath at first, 4) convulsions, 5) jaundice needing treatment, 6) required oxygen, 7) required blood transfusion, 8) Rh incompatibility.

*Alcohol/Tobacco Use During Pregnancy*

Parents self-reported alcohol and/or tobacco use for both prior to knowing of pregnancy, and after knowing of pregnancy. Alcohol/tobacco use during pregnancy was coded as a binary variable indicating self-reported use of either substance, either prior to or after knowing of pregnancy.

*Data Analysis*

Comparisons of differences in raw adiposity markers and global brain measurements by gestational diabetes mellitus (GDM) exposure status were conducted via t-tests (eTable 1). Linear mixed effects models were used to examine associations of GDM exposure with adiposity markers (eFigure 2).

To follow up with significant associations between prenatal diabetes exposure and whole-brain measurements, region of interest (ROI)-based analyses were performed. Cortical gray matter volume of 34 ROIs defined bilaterally on the Desikan-Killany atlas were included in follow-up analyses. Subcortical gray matter volume of 7 ROIs labeled bilaterally by automated segmentation were also included for analysis. Group differences in cortical gray matter volume and subcortical gray matter volume for each ROI were assessed using models predicting bilateral structure based on prenatal exposure to GDM status by hemisphere (right, left) interaction. Family ID, nested within site, was included in the model using random effects; furthermore, subject identification was included as a random effect to pool across hemispheres. Covariates included in the model were the same as for the global brain analysis.

Analyses used linear mixed effects models in R with the *lme4* package. Standardized betas were reported. 95% Wald confidence intervals were calculated based on the local curvature of the likelihood surface. Cohen’s *d* effect sizes were calculated from least-squares means, model residual standard deviation, and residual degrees-of-freedom using the *emmeans* package. *P*-values were calculated using Satterthwaite’s method in the *lmerTest* package. Tests of significance (2-tailed) were corrected for multiple comparisons using the Benjamini-Hochberg false discovery rate (FDR) correction, with *P*< 0.05 as the corrected threshold for significance.

**eResults**

*Results from models with additional adjustment for various covariates*

Additional models were fitted to adjust for: 1) gestational age at birth (eTables 2-4), 2) other health problems during pregnancy (eTables 5-7), 3) health problems at birth (eTables 8-10), and 4) alcohol or tobacco use during pregnancy (eTables 11-13).

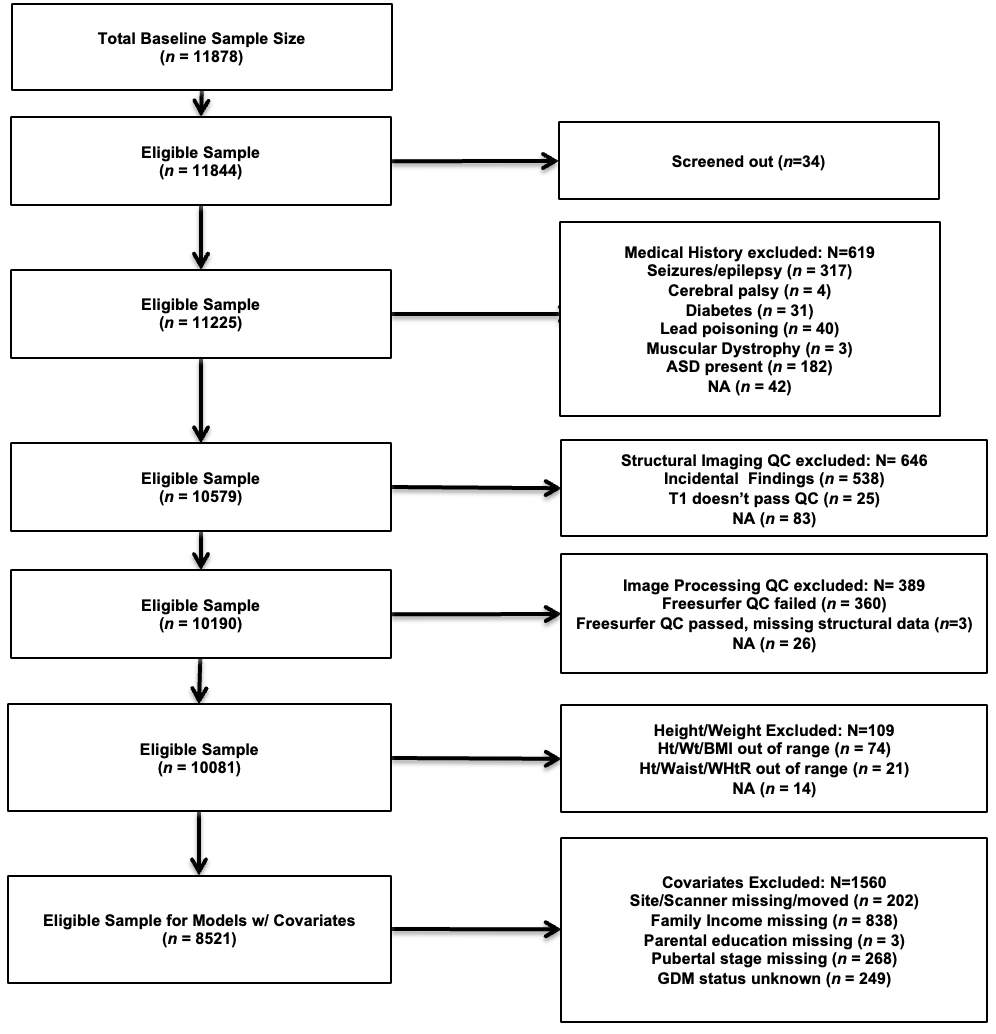

eFigure 1. Sample Exclusion Flow Chart. Flowchart depicts exclusions of ineligible participants based on six categories of criteria: 1) initial screening for eligibility, 2) medical history, including diagnosis of autism spectrum disorder (ASD), 3) quality control (QC) for structural (T1) image and 4) QC of image after processing by Freesurfer software, 5) anthropometric measurements out of range, including for height (Ht), weight (Wt), body mass index (BMI), waist circumference, and waist-to-height ratio (WHtR), and 6) missing data or unknown status for study variables including site and scanner identification, family income, parental education history, pubertal stage assessment, and status of Gestational Diabetes Mellitus (GDM) exposure.

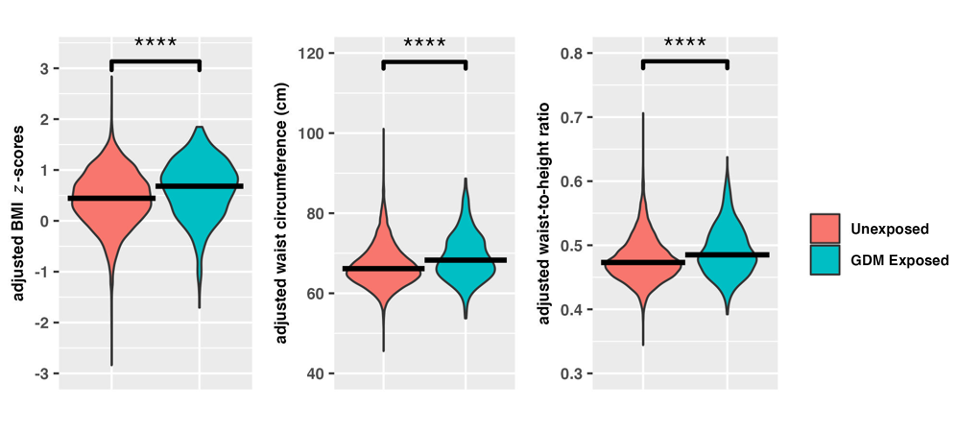

**eFigure 2.** Association of GDM Exposure with Child Adiposity Markers. Violin plots display distributions for adiposity markers (adjusting for family ID nested within site, age, sex, pubertal status, race/ethnicity, family income, highest parental education) separated by prenatal exposure to GDM status. Horizontal bold lines within the violin represent the median of the respective distribution.

**** *P* < 0.0001

eTable 1. Unadjusted Associations of GDM Exposure with Child Adiposity Markers and Global Brain Measurements

|  | **Exposed**  (N=578) | **Unexposed**  (N=7943) |  |
| --- | --- | --- | --- |
|  | Mean (SD) | Mean (SD) | *P*-value |
| BMI *z*-scores | 0.628(1.188) | 0.355(1.129) | **< 0.001** |
| Waist Circumference (cm) | 69.500(11.599) | 66.784(10.339) | **< 0.001** |
| Waist-to-Height Ratio | 0.493(0.073) | 0.475(0.065) | **< 0.001** |
| Total Cortical Surface Area(mm^2^) | 184837.100(18125.450) | 186490.600(17996.370) | **0.003** |
| Mean Cortical Thickness (mm) | 2.767(0.105) | 2.782(0.104) | **0.002** |
| Cortical Gray Matter Volume (mm^3^) | 588800.300(56076.140) | 597699.900(56839.960) | **<0.001** |
| Subcortical Gray Matter Volume (mm^3^) | 59767.460(5120.485) | 60457.280(4963.235) | **0.002** |
| Cerebral White Matter Volume (mm^3^) | 414218.900(50865.290) | 418610.200(48532.490) | **0.045** |

**eTable 2. Associations between GDM Exposure and Child Global Brain Measurements, with Additional Adjustment for Gestational Age**

| **Global Brain Measurements** | **β** | **95% CI** | ***P*-value** | **FDR-adjusted *P*-value** | **Cohen’s *d*** |
| --- | --- | --- | --- | --- | --- |
| Total Cortical Surface Area(mm^2^) | -0.066 | (-0.137, 0.005) | 0.067 | 0.111 | -0.130 |
| Mean Cortical Thickness (mm) | -0.061 | (-0.136, 0.014) | 0.112 | 0.140 | -0.094 |
| Cortical Gray Matter Volume(mm^3^) | -0.045 | (-0.083, -0.007) | 0.022 | 0.108 | -0.141 |
| Subcortical Gray Matter Volume(mm^3^) | -0.045 | (-0.092, 0.002) | 0.061 | 0.111 | -0.120 |
| Cerebral White Matter Volume(mm^3^) | -0.023 | (-0.061, 0.016) | 0.243 | 0.243 | -0.075 |

Analyses completed with adjustment for family ID nested within site, age, sex, puberty status, race/ethnicity, highest parental education, family income, gestational age, scanner model, and handedness. Adjustment for intracranial volume was included for volumetric analyses (*N* = 8483).

**eTable 3. Associations between GDM Exposure and Child Cortical Gray Matter Volume in the Regions of Interest, with Additional Adjustment for Gestational Age**

| **Regional Cortical Gray Matter Volume (mm^3^)** | **β** | **95% CI** | ***P*-value** | **FDR-adjusted *P*-value** | **Cohen’s *d*** |
| --- | --- | --- | --- | --- | --- |
| Banks of the Superior Temporal Sulcus | -0.031 | (-0.090, 0.028) | 0.307 | 0.579 | -0.040 |
| Caudal Anterior Cingulate Cortex | 0.005 | (-0.053, 0.063) | 0.867 | 0.913 | 0.006 |
| Caudal Middle Frontal Gyrus | -0.008 | (-0.070, 0.053) | 0.786 | 0.862 | -0.014 |
| Cuneus Cortex | -0.084 | (-0.146, -0.022) | 0.008 | 0.076 | -0.132 |
| Entorhinal Cortex | 0.054 | (-0.009, 0.117) | 0.095 | 0.318 | 0.071 |
| Fusiform Gyrus | -0.021 | (-0.078, 0.035) | 0.458 | 0.717 | -0.037 |
| Inferior Parietal Cortex | -0.022 | (-0.080, 0.036) | 0.464 | 0.717 | -0.039 |
| Inferior Temporal Gyrus | 0.036 | (-0.021, 0.093) | 0.218 | 0.529 | -0.065 |
| Isthmus Cingulate Cortex | -0.009 | (-0.068, 0.051) | 0.777 | 0.862 | -0.013 |
| Lateral Occipital Cortex | -0.056 | (-0.111, 0.000) | 0.05 | 0.282 | -0.104 |
| Lateral Orbital Frontal Cortex | -0.042 | (-0.098, 0.013) | 0.135 | 0.382 | -0.089 |
| Lingual Gyrus | -0.055 | (-0.121, 0.011) | 0.103 | 0.318 | -0.108 |
| Medial Orbital Frontal Cortex | -0.014 | (-0.071, 0.042) | 0.615 | 0.804 | -0.022 |
| Middle Temporal Gyrus | -0.023 | (-0.077, 0.031) | 0.398 | 0.713 | -0.046 |
| Parahippocampal Gyrus | 0.022 | (-0.041, 0.085) | 0.492 | 0.728 | 0.031 |
| Paracentral Lobule | -0.037 | (-0.100, 0.025) | 0.240 | 0.543 | -0.052 |
| Pars Opercularis | -0.014 | (-0.076, 0.048) | 0.659 | 0.830 | -0.019 |
| Pars Orbitalis | -0.052 | (-0.113, 0.009) | 0.097 | 0.318 | -0.074 |
| Pars Triangularis | -0.012 | (-0.079, 0.055) | 0.725 | 0.862 | -0.017 |
| Pericalcarine Cortex | -0.093 | (-0.163, -0.023) | 0.009 | 0.076 | -0.189 |
| Postcentral Gyrus | -0.032 | (-0.091, 0.028) | 0.297 | 0.579 | -0.056 |
| Posterior Cingulate Cortex | 0.015 | (-0.041, 0.072) | 0.594 | 0.804 | 0.021 |
| Precentral Gyrus | 0.022 | (-0.033, 0.077) | 0.437 | 0.717 | -0.042 |
| Precuneus Cortex | -0.030 | (-0.085, 0.026) | 0.293 | 0.579 | -0.064 |
| Rostral Anterior Cingulate Cortex | -0.009 | (-0.066, 0.048) | 0.761 | 0.862 | -0.012 |
| Rostral Middle Frontal Gyrus | -0.082 | (-0.139, -0.026) | 0.004 | 0.073 | -0.169 |
| Superior Frontal Gyrus | -0.050 | (-0.102, 0.002) | 0.061 | 0.294 | -0.114 |
| Superior Parietal Cortex | -0.040 | (-0.100, 0.020) | 0.189 | 0.495 | -0.078 |
| Superior Temporal Gyrus | -0.098 | (-0.153, -0.042) | 0.001 | **0.019** | -0.194 |
| Supramarginal Gyrus | -0.002 | (-0.058, 0.054) | 0.948 | 0.948 | -0.003 |
| Frontal Pole | -0.005 | (-0.068, 0.059) | 0.886 | 0.913 | -0.006 |
| Temporal Pole | 0.022 | (-0.045, 0.088) | 0.520 | 0.737 | 0.028 |
| Transverse Temporal Cortex | -0.068 | (-0.133, -0.004) | 0.038 | 0.256 | -0.101 |
| Insular Cortex | -0.047 | (-0.102, 0.009) | 0.098 | 0.318 | -0.114 |

Analyses completed with adjustment for family ID nested within site, age, sex, puberty status, race/ethnicity, highest parental education, family income, gestational age, scanner model, handedness, and intracranial volume (*N* = 8483). Exposure by hemisphere interactions were not significant in any ROI after FDR-correction.

**eTable 4. Associations between GDM Exposure and Child Global Brain Measurements, with Additional Adjustment for Health Problems During Pregnancy**

| **Global Brain Measurements** | **β** | **95% CI** | ***P*-value** | **FDR-adjusted *P*-value** | **Cohen’s *d*** |
| --- | --- | --- | --- | --- | --- |
| Total Cortical Surface Area(mm^2^) | -0.065 | (-0.136, -0.005) | 0.069 | 0.116 | -0.128 |
| Mean Cortical Thickness(mm) | -0.059 | (-0.134, 0.015) | 0.120 | 0.151 | -0.092 |
| Cortical Gray Matter Volume(mm^3^) | -0.047 | (-0.086, -0.009) | 0.015 | 0.073 | -0.150 |
| Subcortical Gray Matter Volume(mm^3^) | -0.045 | (-0.092, 0.002) | 0.061 | 0.116 | -0.126 |
| Cerebral White Matter Volume(mm^3^) | -0.025 | (-0.064, 0.013) | 0.196 | 0.196 | -0.083 |

Analyses completed with adjustment for family ID nested within site, age, sex, puberty status, race/ethnicity, highest parental education, family income, health problems during pregnancy, scanner model, and handedness. Adjustment for intracranial volume was included for volumetric analyses (*N* = 8521).

**eTable 5. Associations between GDM Exposure and Child Cortical Gray Matter Volume in the Regions of Interests, with Additional Adjustment for Health Problems During Pregnancy**

| **Regional Cortical Gray Matter Volume (mm^3^)** | **β** | **95% CI** | ***P*-value** | **FDR-adjusted *P*-value** | **Cohen’s *d*** |
| --- | --- | --- | --- | --- | --- |
| Banks of the Superior Temporal Sulcus | -0.036 | (-0.096, 0.023) | 0.231 | 0.466 | -0.047 |
| Caudal Anterior Cingulate Cortex | 0.006 | (-0.052, 0.063) | 0.847 | 0.847 | 0.006 |
| Caudal Middle Frontal Gyrus | -0.014 | (-0.075, 0.047) | 0.647 | 0.759 | -0.023 |
| Cuneus Cortex | -0.079 | (-0.141, -0.018) | 0.012 | 0.111 | -0.125 |
| Entorhinal Cortex | 0.056 | (-0.007, 0.119) | 0.083 | 0.282 | 0.074 |
| Fusiform Gyrus | -0.027 | (-0.083, 0.029) | 0.345 | 0.580 | -0.047 |
| Inferior Parietal Cortex | -0.027 | (-0.085, 0.031) | 0.358 | 0.580 | -0.049 |
| Inferior Temporal Gyrus | 0.037 | (-0.019, 0.094) | 0.196 | 0.466 | 0.068 |
| Isthmus Cingulate Cortex | -0.009 | (-0.068, 0.050) | 0.772 | 0.811 | -0.013 |
| Lateral Occipital Cortex | -0.050 | (-0.106, 0.076) | 0.076 | 0.282 | -0.094 |
| Lateral Orbital Frontal Cortex | -0.043 | (-0.099, 0.012) | 0.012 | 0.124 | -0.091 |
| Lingual Gyrus | -0.059 | (-0.124, 0.007) | 0.007 | 0.282 | -0.115 |
| Medial Orbital Frontal Cortex | -0.036 | (-0.069, 0.043) | 0.643 | 0.759 | -0.020 |
| Middle Temporal Gyrus | -0.036 | (-0.090, 0.018) | 0.192 | 0.466 | -0.071 |
| Parahippocampal Gyrus | 0.020 | (-0.043, 0.083) | 0.534 | 0.759 | 0.028 |
| Paracentral Lobule | -0.039 | (-0.101, 0.023) | 0.220 | 0.466 | -0.054 |
| Pars Opercularis | -0.016 | (-0.079, 0.046) | 0.612 | 0.759 | -0.022 |
| Pars Orbitalis | -0.056 | (-0.117, -0.005) | 0.071 | 0.282 | -0.080 |
| Pars Triangularis | -0.017 | (-0.084, 0.050) | 0.614 | 0.759 | -0.024 |
| Pericalcarine Cortex | -0.088 | (-0.158, -0.019) | 0.013 | 0.111 | -0.179 |
| Postcentral Gyrus | -0.033 | (-0.092, 0.026) | 0.277 | 0.524 | -0.058 |
| Posterior Cingulate Cortex | 0.013 | (-0.043, 0.069) | 0.647 | 0.759 | 0.018 |
| Precentral Gyrus | 0.022 | (-0.033, 0.026) | 0.429 | 0.663 | 0.042 |
| Precuneus Cortex | -0.030 | (-0.085, 0.026) | 0.293 | 0.525 | -0.064 |
| Rostral Anterior Cingulate Cortex | -0.012 | (-0.069, 0.045) | 0.679 | 0.770 | -0.016 |
| Rostral Middle Frontal Gyrus | -0.085 | (-0.142, -0.029) | 0.003 | 0.052 | -0.175 |
| Superior Frontal Gyrus | -0.053 | (-0.105, -0.001) | 0.046 | 0.258 | -0.121 |
| Superior Parietal Cortex | -0.036 | (-0.096, 0.023) | 0.233 | 0.466 | -0.071 |
| Superior Temporal Gyrus | -0.099 | (-0.155, -0.044) | <0.001 | **0.015** | -0.197 |
| Supramarginal Gyrus | -0.009 | (-0.064, 0.047) | 0.761 | 0.811 | -0.014 |
| Frontal Pole | -0.009 | (-0.072, 0.055) | 0.788 | 0.811 | -0.011 |
| Temporal Pole | 0.020 | (-0.046, 0.086) | 0.557 | 0.759 | 0.025 |
| Transverse Temporal Cortex | -0.073 | (-0.137, -0.009) | 0.026 | 0.174 | -0.108 |
| Insular Cortex | -0.047 | (-0.102, 0.008) | 0.096 | 0.296 | 0.296 |

Analyses completed with adjustment for family ID nested within site, age, sex, puberty status, race/ethnicity, highest parental education, family income, health problems during pregnancy scanner model, handedness, and intracranial volume (*N* = 8521). Exposure by hemisphere interactions were not significant in any ROI after FDR-correction.

**eTable 6. Associations between GDM Exposure and Child Global Brain Measurements, with Additional Adjustment for Health Problems at Birth**

| **Global Brain Measurements** | **β** | **95% CI** | ***P*-value** | **FDR-adjusted *P*-value** | **Cohen’s *d*** |
| --- | --- | --- | --- | --- | --- |
| Total Cortical Surface Area(mm^2^) | -0.071 | (-0.141, -0.000) | 0.049 | 0.91 | -0.139 |
| Mean Cortical Thickness(mm) | -0.061 | (-0.135, 0.014) | 0.111 | 0.140 | -0.094 |
| Cortical Gray Matter Volume(mm^3^) | -0.049 | (-0.088, -0.011) | 0.011 | 0.055 | -0.156 |
| Subcortical Gray Matter Volume(mm^3^) | -0.046 | (-0.093, 0.001) | 0.055 | 0.091 | -0.076 |
| Cerebral White Matter Volume(mm^3^) | -0.023 | (-0.061, 0.015) | 0.238 | 0.238 | -0.123 |

Analyses completed with adjustment for family ID nested within site, age, sex, puberty status, race/ethnicity, highest parental education, family income, health problems at birth, scanner model, and handedness. Adjustment for intracranial volume was included for volumetric analyses. Adjustment for intracranial volume was included for volumetric analyses (*N* = 8515).

**eTable 7. Associations between GDM Exposure and Child Cortical Gray Matter Volume in the Regions of Interests, with Additional Adjustment for Health Problems at Birth**

| **Regional Cortical Gray Matter Volume (mm^3^)** | **β** | **95% CI** | ***P*-value** | **FDR-adjusted *P*-value** | **Cohen’s *d*** |
| --- | --- | --- | --- | --- | --- |
| Banks of the Superior Temporal Sulcus | -0.035 | (-0.094, 0.024) | 0.0249 | 0.445 | -0.045 |
| Caudal Anterior Cingulate Cortex | 0.006 | (-0.051, 0.064) | 0.829 | 0.854 | 0.007 |
| Caudal Middle Frontal Gyrus | -0.014 | (-0.075, 0.047) | 0.643 | 0.769 | -0.023 |
| Cuneus Cortex | -0.078 | (-0.140, -0.017) | 0.013 | 0.127 | -0.124 |
| Entorhinal Cortex | 0.059 | (-0.004, 0.122) | 0.068 | 0.230 | 0.078 |
| Fusiform Gyrus | -0.026 | (-0.082, 0.031) | 0.373 | 0.603 | -0.044 |
| Inferior Parietal Cortex | -0.029 | (-0.086, 0.029) | 0.334 | 0.568 | -0.051 |
| Inferior Temporal Gyrus | 0.036 | (-0.020, 0.093) | 0.211 | 0.435 | 0.065 |
| Isthmus Cingulate Cortex | -0.013 | (-0.073, 0.046) | 0.658 | 0.769 | -0.018 |
| Lateral Occipital Cortex | -0.052 | (-0.108, 0.004) | 0.067 | 0.230 | -0.097 |
| Lateral Orbital Frontal Cortex | -0.047 | (-0.102, 0.009) | 0.098 | 0.277 | -0.098 |
| Lingual Gyrus | -0.060 | (-0.126, 0.006) | 0.074 | 0.230 | -0.118 |
| Medial Orbital Frontal Cortex | -0.014 | (-0.070, 0.043) | 0.635 | 0.769 | -0.023 |
| Middle Temporal Gyrus | -0.037 | (-0.091, 0.017) | 0.183 | 0.414 | -0.072 |
| Parahippocampal Gyrus | 0.026 | (-0.037, 0.089) | 0.414 | 0.639 | 0.036 |
| Paracentral Lobule | -0.039 | (-0.101, 0.023) | 0.220 | 0.435 | -0.054 |
| Pars Opercularis | -0.020 | (-0.083, 0.042) | 0.522 | 0.710 | -0.028 |
| Pars Orbitalis | -0.058 | (-0.119, 0.002) | 0.060 | 0.230 | -0.084 |
| Pars Triangularis | -0.019 | (-0.086, 0.047) | 0.567 | 0.742 | -0.028 |
| Pericalcarine Cortex | -0.087 | (-0.156, -0.017) | 0.015 | 0.127 | -0.176 |
| Postcentral Gyrus | -0.036 | (-0.096, 0.023) | 0.230 | 0.435 | -0.064 |
| Posterior Cingulate Cortex | 0.012 | (-0.044, 0.068) | 0.679 | 0.769 | 0.016 |
| Precentral Gyrus | 0.022 | (-0.034, 0.077) | 0.444 | 0.656 | 0.041 |
| Precuneus Cortex | -0.038 | (-0.093, 0.017) | 0.174 | 0.414 | -0.082 |
| Rostral Anterior Cingulate Cortex | -0.010 | (-0.067, 0.046) | 0.720 | 0.790 | -0.014 |
| Rostral Middle Frontal Gyrus | -0.084 | (-0.141, -0.028) | 0.003 | 0.056 | -0.174 |
| Superior Frontal Gyrus | -0.051 | (-0.104, 0.001) | 0.054 | 0.230 | -0.117 |
| Superior Parietal Cortex | -0.046 | (-0.106, 0.014) | 0.135 | 0.353 | -0.089 |
| Superior Temporal Gyrus | -0.099 | (-0.154, -0.043) | <0.001 | **0.016** | -0.196 |
| Supramarginal Gyrus | -0.008 | (-0.064, 0.047) | 0.765 | 0.813 | -0.014 |
| Frontal Pole | -0.006 | (-0.069, 0.057) | 0.859 | 0.859 | -0.008 |
| Temporal Pole | 0.024 | (-0.043, 0.090) | 0.482 | 0.683 | 0.030 |
| Transverse Temporal Cortex | -0.075 | (-0.140, -0.011) | 0.021 | 0.145 | -0.112 |
| Insular Cortex | -0.051 | (-0.106, 0.005) | 0.072 | 0.230 | -0.124 |

Analyses completed with adjustment for family ID nested within site, age, sex, puberty status, race/ethnicity, highest parental education, family income, health problems at birth, scanner model, handedness, and intracranial volume (*N* = 8515). Exposure by hemisphere interactions were not significant in any ROI after FDR-correction.

**eTable 8. Associations between GDM Exposure and Child Global Brain Measurements, with Additional Adjustment for Alcohol or Tobacco Use During Pregnancy**

| **Global Brain Measurements** | **β** | **95% CI** | ***P*-value** | **FDR-adjusted *P*-value** | **Cohen’s *d*** |
| --- | --- | --- | --- | --- | --- |
| Total Cortical Surface Area(mm^2^) | -0.065 | (-0.136, 0.006) | 0.075 | 0.124 | -0.127 |
| Mean Cortical Thickness(mm) | -0.060 | (-0.135, 0.016) | 0.122 | 0.152 | -0.092 |
| Cortical Gray Matter Volume(mm^3^) | -0.049 | (-0.087, -0.010) | 0.013 | 0.064 | -0.153 |
| Subcortical Gray Matter Volume(mm^3^) | -0.051 | (-0.099, -0.004) | 0.034 | 0.085 | -0.138 |
| Cerebral White Matter Volume(mm^3^) | -0.026 | (-0.065, 0.012) | 0.183 | 0.183 | -0.087 |

Analyses completed with adjustment for family ID nested within site, age, sex, puberty status, race/ethnicity, highest parental education, family income, alcohol or tobacco use during pregnancy, scanner model, and handedness. Adjustment for intracranial volume was included for volumetric analyses (*N* = 8184).

**eTable 9. Associations between GDM Exposure and Child Cortical Gray Matter Volume in the Regions of Interests, with Additional Adjustment for Alcohol or Tobacco Use During Pregnancy**

| **Regional Cortical Gray Matter Volume (mm^3^)** | **β** | **95% CI** | ***P*-value** | **FDR-adjusted *P*-value** | **Cohen’s *d*** |
| --- | --- | --- | --- | --- | --- |
| Banks of the Superior Temporal Sulcus | -0.037 | (-0.097, 0.023) | 0.226 | 0.453 | -0.048 |
| Caudal Anterior Cingulate Cortex | 0.000 | (-0.058, 0.059) | 0.994 | 0.994 | 0.000 |
| Caudal Middle Frontal Gyrus | -0.011 | (-0.072, 0.051) | 0.735 | 0.821 | -0.017 |
| Cuneus Cortex | -0.085 | (-0.148, -0.022) | 0.008 | 0.088 | -0.134 |
| Entorhinal Cortex | 0.062 | (-0.002, 0.125) | 0.058 | 0.198 | 0.081 |
| Fusiform Gyrus | -0.025 | (-0.082, 0.032) | 0.391 | 0.635 | -0.043 |
| Inferior Parietal Cortex | -0.027 | (-0.085, 0.032) | 0.371 | 0.635 | -0.048 |
| Inferior Temporal Gyrus | 0.041 | (-0.017, 0.098) | 0.164 | 0.381 | 0.074 |
| Isthmus Cingulate Cortex | -0.013 | (-0.072, 0.047) | 0.678 | 0.794 | -0.019 |
| Lateral Occipital Cortex | -0.056 | (-0.112, 0.000) | 0.051 | 0.198 | -0.104 |
| Lateral Orbital Frontal Cortex | -0.048 | (-0.104, 0.008) | 0.093 | 0.288 | -0.100 |
| Lingual Gyrus | -0.054 | (-0.120, 0.013) | 0.112 | 0.317 | -0.106 |
| Medial Orbital Frontal Cortex | -0.016 | (-0.072, 0.041) | 0.591 | 0.717 | -0.024 |
| Middle Temporal Gyrus | -0.038 | (-0.093, 0.016) | 0.168 | 0.381 | -0.075 |
| Parahippocampal Gyrus | 0.024 | (-0.039, 0.088) | 0.452 | 0.673 | 0.034 |
| Paracentral Lobule | -0.048 | (-0.110, 0.015) | 0.133 | 0.349 | -0.066 |
| Pars Opercularis | -0.0018 | (-0.081, 0.045) | 0.570 | 0.717 | -0.027 |
| Pars Orbitalis | -0.059 | (-0.121, 0.002) | 0.058 | 0.198 | -0.085 |
| Pars Triangularis | -0.020 | (-0.087, 0.048) | 0.568 | 0.717 | -0.027 |
| Pericalcarine Cortex | -0.091 | (-0.161, -0.021) | 0.011 | 0.094 | -0.185 |
| Postcentral Gyrus | -0.039 | (-0.099, 0.021) | 0.202 | 0.430 | -0.069 |
| Posterior Cingulate Cortex | -0.000 | (-0.057, 0.056) | 0.988 | 0.994 | -0.001 |
| Precentral Gyrus | 0.019 | (-0.037, 0.075) | 0.502 | 0.682 | 0.036 |
| Precuneus Cortex | -0.024 | (-0.080, 0.031) | 0.392 | 0.635 | -0.052 |
| Rostral Anterior Cingulate Cortex | -0.024 | (-0.079, 0.035) | 0.455 | 0.673 | -0.029 |
| Rostral Middle Frontal Gyrus | -0.085 | (-0.142, -0.028) | 0.003 | 0.059 | -0.174 |
| Superior Frontal Gyrus | -0.053 | (-0.105, 0.000) | 0.051 | 0.198 | -0.120 |
| Superior Parietal Cortex | -0.034 | (-0.094, 0.027) | 0.273 | 0.515 | -0.066 |
| Superior Temporal Gyrus | -0.101 | (-0.158, -0.046) | <0.001 | **0.013** | -0.202 |
| Supramarginal Gyrus | -0.006 | (-0.062, 0.051) | 0.845 | 0.898 | -0.007 |
| Frontal Pole | -0.010 | (-0.074, 0.054) | 0.749 | 0.821 | -0.014 |
| Temporal Pole | 0.024 | (-0.043, 0.091) | 0.478 | 0.678 | 0.031 |
| Transverse Temporal Cortex | -0.080 | (-0.145, -0.015) | 0.016 | 0.107 | -0.118 |
| Insular Cortex | -0.057 | (-0.113, -0.001) | 0.045 | 0.198 | -0.139 |

Analyses completed with adjustment for family ID nested within site, age, sex, puberty status, race/ethnicity, highest parental education, family income, alcohol or tobacco use during pregnancy, scanner model, handedness, and intracranial volume (*N* = 8184). Exposure by hemisphere interactions were not significant in any ROI after FDR-correction.

**eTable 10. Associations between GDM Exposure and Child Subcortical Gray Matter Volume in the Regions of Interests**

| **Regional Subcortical Gray Matter Volume (mm^3^)** | **β** | **95% CI** | ***P*-value** | **FDR-adjusted *P*-value** | **Cohen’s *d*** |
| --- | --- | --- | --- | --- | --- |
| Thalamus | -0.047 | (-0.099, 0.004) | 0.069 | 0.196 | -0.135 |
| Caudate | -0.030 | (-0.099, 0.039) | 0.389 | 0.680 | -0.108 |
| Putamen | -0.084 | (-0.151, -0.017) | 0.014 | 0.099 | -0.235 |
| Pallidum | -0.007 | (-0.066, 0.053) | 0.826 | 0.826 | -0.009 |
| Hippocampus | -0.019 | (-0.080, 0.042) | 0.543 | 0.760 | -0.044 |
| Amygdala | -0.050 | (-0.107, 0.007) | 0.084 | 0.196 | -0.096 |
| Accumbens Area | -0.008 | (-0.069, 0.052) | 0.789 | 0.826 | -0.014 |

Analyses completed with adjustment for family ID nested within site, age, sex, puberty status, race/ethnicity, highest parental education, family income, scanner model, handedness, and intracranial volume.

Exposure by hemisphere interactions were not significant in any ROI after FDR-correction.

**eTable 11.** **Mediation Results of Gestational Diabetes Mellitus Status Exposure (GDM), Cortical Gray Matter Volume (GMV), and Adiposity Measures.**

|  | **GDM** $\boldsymbol{\to}$ **Cortical GMV** $\boldsymbol{\to}$  **BMI *z*-scores** | | **GDM** $\boldsymbol{\to}$ **Cortical GMV** $\boldsymbol{\to}$  **Waist Circumference** | | **GDM** $\boldsymbol{\to}$**Cortical GMV** $\boldsymbol{\to}$  **Waist-Height Ratio** | |
| --- | --- | --- | --- | --- | --- | --- |
|  | **Estimate**  **(95% CI)** | ***P*-value** | **Estimate**  **(95% CI)** | ***P*-value** | **Estimate**  **(95% CI)** | ***P*-value** |
| Total Effect*  (Y~X) | 0.168  (0.077, 0.255) | **<0.001** | 0.198  (0.103, 0.295) | **<0.001** | 0.195  (0.107, 0.290) | **<0.001** |
| Direct Effect*  (Y~X+M) | 0.161  (0.072, 0.247) | **<0.001** | 0.190  (0.098, 0.287) | **<0.001** | 0.191  (0.107, 0.285) | **<0.001** |
| Mediated Effect* | 0.007  (0.002, 0.013) | **0.012** | 0.008  (0.002, 0.015) | **0.012** | 0.005  (0.001, 0.009) | **0.012** |
| Proportion Mediated | 0.040  (0.010, 0.096) | **0.012** | 0.040  (0.010, 0.089) | **0.012** | 0.024  (0.006, 0.057) | **0.012** |

*Total Effect and Direct Effect are standardized regression coefficients of X (GDM) on Y (adiposity measures) without and with the addition of mediator M (Cortical GMV). The mediated effect indicates the amount of mediation attributed to M.
